# Pattern, preferences, barriers, and correlates of self-reported physical activity in adults with borderline personality disorder: An online survey in western countries

**DOI:** 10.1101/2022.05.24.22275513

**Authors:** Samuel St-Amour, Lionel Cailhol, Josyanne Lapointe, Déborah Ducasse, Gabrielle Landry, Paquito Bernard

## Abstract

Borderline personality disorder (BPD) is characterized by an instability of self-image, interpersonal relationships, and emotions and is highly comorbid with other disorders. Physical activity has shown great results in treating these disorders. Physical activity intervention should be built considering the preferences and barriers of the targeted individuals. However, to this day no study analyzed the preferences and barriers to physical activity in individuals with BPD, which is the goal of this study. We used an online survey to question 192 adults with a self-reported diagnosis of BPD from Canada, France, the United States, England, Switzerland, and New Zealand. Participants complete 607 minutes of physical activity weekly on average. They prefer walking (66.7%), biking (33.3%), aquatic activities (29.0%), and running (24.2%). Their main barriers to physical activity are having a friend over, having other engagements, and recovering from an injury. They also prefer doing individual supervised physical activity outside and in a long session of moderate intensity. Finally, a majority of participants are interested in receiving physical activity advice, but most did not. The professionals from whom they would prefer to receive advice are trainers, psychiatrists, physical therapists, and psychologists. These results are important to better tailor future physical activity interventions for adults with BPD.

## 1. Introduction

Borderline personality disorder (BPD) is the fourth most prevalent personality disorder in general population and the most prevalent in clinical settings (Gunderson et al., 2018). It is characterized by an instability of the self, individual goals, interpersonal relationships and affects (Gunderson et al., 2018). The biosocial development model of BPD suggests that emotion dysregulation is among the core components of the disorder and underlies some of its characteristic behaviours (Crowell et al., 2009). Moreover, emotion dysregulation has been linked to a lower quality of life and daily functioning and a poorer therapeutic relationship (Gunderson et al., 2018). Individuals with BPD are also highly at risk to commit suicide with 83% having a history of suicide attempt (Soloff et al., 2002) and 8 to 10% dying from suicide (Biskin, 2015).

Those individuals also frequently present comorbid physical and mental disorders (El-Gabalawy et al., 2010; Shah & Zanarini, 2018). Mood disorders are the most frequent mental comorbid disorders in individuals with BPD with a lifetime prevalence of 96% (Shah & Zanarini, 2018). The main physical comorbid disorders in individuals with BPD include obesity, cardiovascular diseases and diabetes with one-year prevalence of 34%, 15%, and 9% respectively (Castle, 2019). Moreover, cardiovascular and metabolic disorders are among the greatest mortality causes with over 20% of death in this population (Cailhol et al., 2017; Kuo et al., 2019).

Among the treatments to address these different disorders, physical activity (PA) has been linked to improvements in many of their components and symptoms. With a lifetime prevalence of 96%, mood disorders are the most prevalent comorbid disorder in adults with BPD (Shah & Zanarini, 2018). The last Canadian treatment guidelines for mood disorders included PA as mono- or adjunct therapy for every level of depression severity with the strongest level of proof (Ravindran et al., 2016). Finally, PA was also found efficient in preventing and reducing risk factors of obesity, cardiovascular diseases, and diabetes in individuals with mental disorder (Vancampfort et al., 2013). However, only one study to our knowledge analyzed the short-term effect of PA on BPD itself (St-Amour et al., 2022). No study examined the barriers, preferences and pattern of PA in adults with BPD as reported in a recent review (St-Amour, Cailhol, et al., 2021; St-Amour et al., 2023).

To efficiently study PA’s effects in individuals with BPD we can learn from the previous research done with individuals with other mental disorders. Among the many challenges of studying PA effects in individuals with mental disorders, dropout and adherence are among the greatest (Stubbs & Rosenbaum, 2018). Indeed, dropout rates in studies analyzing PA’s effect in individuals with mental disorders may be as high as 90% and are heavily influenced by study and intervention characteristics (Firth et al., 2015). Moreover, participants who completed those studies did not necessarily attend all sessions. Attendance widely varied in these studies ranging from 30% to 100% of the planned seances (Firth et al., 2015). It is therefore important to provide optimal conditions to reach and more importantly keep participants in future PA studies. The latest guidelines for PA interventions in individuals with mental disorders suggest tailoring those interventions in light of the preferences and barriers of the targeted individuals or population (Romain & Bernard, 2018). It is therefore important to identify those preferences and barriers in individuals with BPD, which has not yet been done to our knowledge, to plan effective interventions. Since BPD is a more relational disorder than other mental disorders in its nature (Gunderson et al., 2018), preferences regarding PA is done and main barriers preventing PA participation could differ from populations with other mental disorders.

There is a growing interest in research for PA in individuals with BPD. However, to our knowledge there are very few observational studies (St-Amour et al., 2023) and only one experimental study (St-Amour et al., 2022) addressing this topic. To develop well-adapted PA interventions for adults with BPD in the future, we need to identify the most important factors associated with adherence.

Being the first study in this population, the aims are to: 1-describe the self-reported PA level of adults with BPD; 2-describe the main preferences regarding type, location, intensity, supervision, and advice related to PA in adults with BPD; 3-describe the main barriers to practise PA in adults with BPD; and 4-examine the main sociodemographic and health variables associated to the self-reported level of PA in adults with BPD.

## 2. Methods

This online cross-sectional study in western countries was performed using the LimeSurvey platform hosted on the Université du Québec à Montréal’s servers. The survey was promoted online in Canada, France, England, the United States, New Zealand, Switzerland, Belgium, and Australia, where the research team had research collaborators, with YouTube videos, a Facebook page and posts on Facebook groups, forums, and chat groups dedicated to adults with BPD (with the permission of the respective administrators). Psychiatrists in France and Canada helped promote this survey by sharing it with their patients, their colleagues, and in different networks regrouping patients and professionals working with patients with BPD. To be included, participants had to report: 1-being at least 18 years old, 2-living in Canada, France, England, the United States, New Zealand, Switzerland, Belgium, or Australia, and 3-having received a BPD diagnosis from a healthcare professional. All participants had to read and agree to the online information and consent form and were given the opportunity to contact the research team prior to filling-out the survey. They were warned that the survey may address sensitive topics, and local help resources were given to participants according to their reported country of residence at the bottom of each page (Batterham, 2014; Choi et al., 2017). This study has been approved by the ethics boards from the Eastern Montreal Integrated University Health and Social Services Centres (2021-2330) and the Université du Québec à Montréal (3997_e_2021).

### 2.1 Questionnaires

Every questionnaire used in this survey has been previously validated in English and in French. All items and questionnaires in English and French are available in open access (https://osf.io/5u6am/). Questionnaires included in this survey relate to PA habits and the main correlates of PA found in the literature (income, education level, disorder severity and duration, medication use, substance use, and suicide ideations; St-Amour, Hains-Monfette, et al., 2021; Vancampfort et al., 2012, 2018).

#### 2.1.1. Sociodemographic characteristics

The following sociodemographic characteristics have been collected: country of residence, sex at birth, age, education level, marital status, height, weight, and household income. Participants have also been asked about their psychiatric follow-up duration, their psychotropic medication use, their comorbid disorders and the numbers of past mental disorder hospitalization and suicide attempts. Afterward, they had to rate their social status with the MacArthur Scale (Adler et al., 2008).

#### 2.1.2. Clinical characteristics

Participants then filled out the *Borderline Symptoms List-Short Form* (BSL-23) validated to measure the presence and severity of symptoms attributed to BPD (Bohus et al., 2009). Then, they filled the *Beck Depression Inventory-Short Form* (BDI-SF) validated to measure the clinical depression risk in adults with BPD. A score of 10 and higher indicate the presence of depression (Furlanetto et al., 2005). Difficulties in emotion regulation have been assessed with the *Difficulties in Emotion Regulation Scale-Short Form* (DERS-18) that have been validated in adults with BPD (Victor & Klonsky, 2016).

#### 2.1.3. Health behaviours

Substance use disorders have been assessed with the *Alcohol, Smoking and Substance Involvement Screening Test* (ASSIST). This questionnaire developed and validated by the World Health Organization (WHO) measures substance use disorders related to tobacco, alcohol, cannabis, cocaine, amphetamines, inhalants, sedatives, hallucinogens, and opioids. A score is obtained for each substance by adding the score from each question. A score of 4 and more (11 and more for alcohol) indicate a moderated substance use disorder and a score of 27 and more (including alcohol) indicate a severe substance use disorder (WHO ASSIST Workin Group, 2002). Participants’ insomnia was assessed with the *Insomnia Severity Index* (ISI) in which a score higher than 15 indicating a risk of clinical insomnia (Bastien et al., 2001). PA was assessed using the *Global Physical Activity Questionnaire* (GPAQ) developed by the WHO to measure work, leisure, travel, and total PA (Armstrong & Bull, 2006).

#### 2.1.4. Physical activity preferences and barriers

Preferences regarding type, intensity, context, supervision, and advice of PA and main barriers to PA have been assessed using questionnaires from previous studies in individuals with mental disorders (Abrantes et al., 2011; Romain, Longpré-Poirier, et al., 2020).

### 2.2. Statistical analysis

Descriptive statistics (N, %, mean) are used to describe the sociodemographic characteristics, the substance use disorder, insomnia, and PA level, preferences, and barriers in this sample. A Poisson multivariate regression was done to identify the main sociodemographic and clinical factors associated with weekly PA level. The age, social level, education level, household income, body mass index, emotion regulation difficulties, BPD symptoms, tobacco and alcohol use disorders, depression, past suicide attempts, psychotropic medication use and self-efficacy were included in the model. These independent variables were selected based on previous papers examining the determinants of PA in adults with mental disorders (Schuch et al., 2017; Vancampfort et al., 2012, 2015). A stepwise variable selection in which variables were added if they reduced model Akaike information criterion was carried out. The multicollinearity of this model has been tested by calculating the variance inflation factor (VIF).

No predictors had VIF values greater than 10 after the stepwise variable selection procedure. Statistical analyses were done with the R software version 4.2 with the libraries “ggstaplot”, “stargazer”, “bestglm”, and “summarytools” (Patil, 2021; Zhang, 2016). Research materials, data and R codes are available in open access on *Open Science Framework* (https://osf.io/c3gvx/).

## 3. Results

The online survey was completed by 288 participants who reported having a BPD diagnosis, but only 192 filled out the survey at least up to the GPAQ providing PA data. Therefore, the results are based on this sample of 192 that completed the GPAQ. Description of the sample is presented in Table 1.

**Table 1:** Descriptive data of subjects.

|  | N (%) |
| --- | --- |
| Sex at birth |  |
| (Women) | 164 (85.9) |
| <35 years | 111 (57.8) |
| < University education | 118 (61.5) |
| Subjective Social Status M (SD) | 4.5 (2.0) |
| Country of residence |  |
| Canada | 96 (50.0) |
| France | 78 (40.6) |
| United States | 9 (4.7) |
| England | 7 (3.6) |
| Switzerland | 2 (1.0) |
| Income |  |
| <20,000 | 71 (37.0) |
| 20,000-39,999 | 36 (18.8) |
| 40,000-59,999 | 21 (10.9) |
| 60,000-79,999 | 24 (12.5) |
| 80,000-99,999 | 10 (5.2) |
| ≥100,000 | 8 (4.2) |
| Don't know | 12 (6.2) |
| No response | 10 (5.2) |
| Marital status |  |
| Single | 95 (47.4) |
| Civil union | 49 (25.5) |
| Married | 23 (12.0) |
| Divorced | 19 (9.9) |
| Widowed | 2 (1.0) |
| Other | 4 (2.1) |
| Body mass index M (SD) | 26.4 (8.1) |
| Self-reported diagnosis of physical disease | 42 (21.9) |
| Self-reported diagnosis of mental disorder | 101 (52.6) |
| Psychotropic medication | 140 (72.9) |
| Currently in psychotherapy | 95 (47.4) |
| Have been hospitalized | 113 (58.9) |
| Attempted suicide | 126 (65.6) |
| Total substance involvement |  |
| High | 111 (57.8) |
| Moderate | 20 (10.4) |
| None | 61 (31.8) |
| Insomnia severity index M (SD) | 13.2 (6.5) |
| Borderline symptoms list-short form M (SD) | 1.9 (0.9) |
| Beck Depression inventory | 16.0 (7.7) |
| Difficulty in emotion regulation scale M (SD) | 59.9(13.5) |

### 3.1. Levels of PA

In this sample, 122 (65.9%) participants declared being physically active according to the guidelines from the WHO (completing at least 150 minutes of PA weekly; WHO. Regional Office for Europe & United Nations Economic Commission for Europe, 2022). On average, participants complete 165.5 (SD = 282.1) minutes of travel-related PA, 315.2 (SD = 685.7) minutes of work-related PA, and 126.0 (SD = 224.9) minutes of leisure-related PA for a total of 606.7 (SD = 766.2) minutes of PA weekly including all three domains. In this sample, men performed significantly more PA than women (p = 0.002). For detailed results comparing total PA levels according to body mass index, sex, level of education, age, and country, see supplementary material (Figure S1).

### 3.2. Barriers

Barriers preventing participants from engaging in PA were measured with self-efficacy scales from 0 to 100. For each barrier, participants had to rate their self-efficacy to engage in PA when encountering said barrier. Scores were reverse for clarity, therefore a higher score means a more important barrier. The PA barriers are illustrated in Figure 1. For detailed barriers according to age, education level, sex, body mass index, and country, see supplementary material (Figures S2-S6).

**Figure 1:**
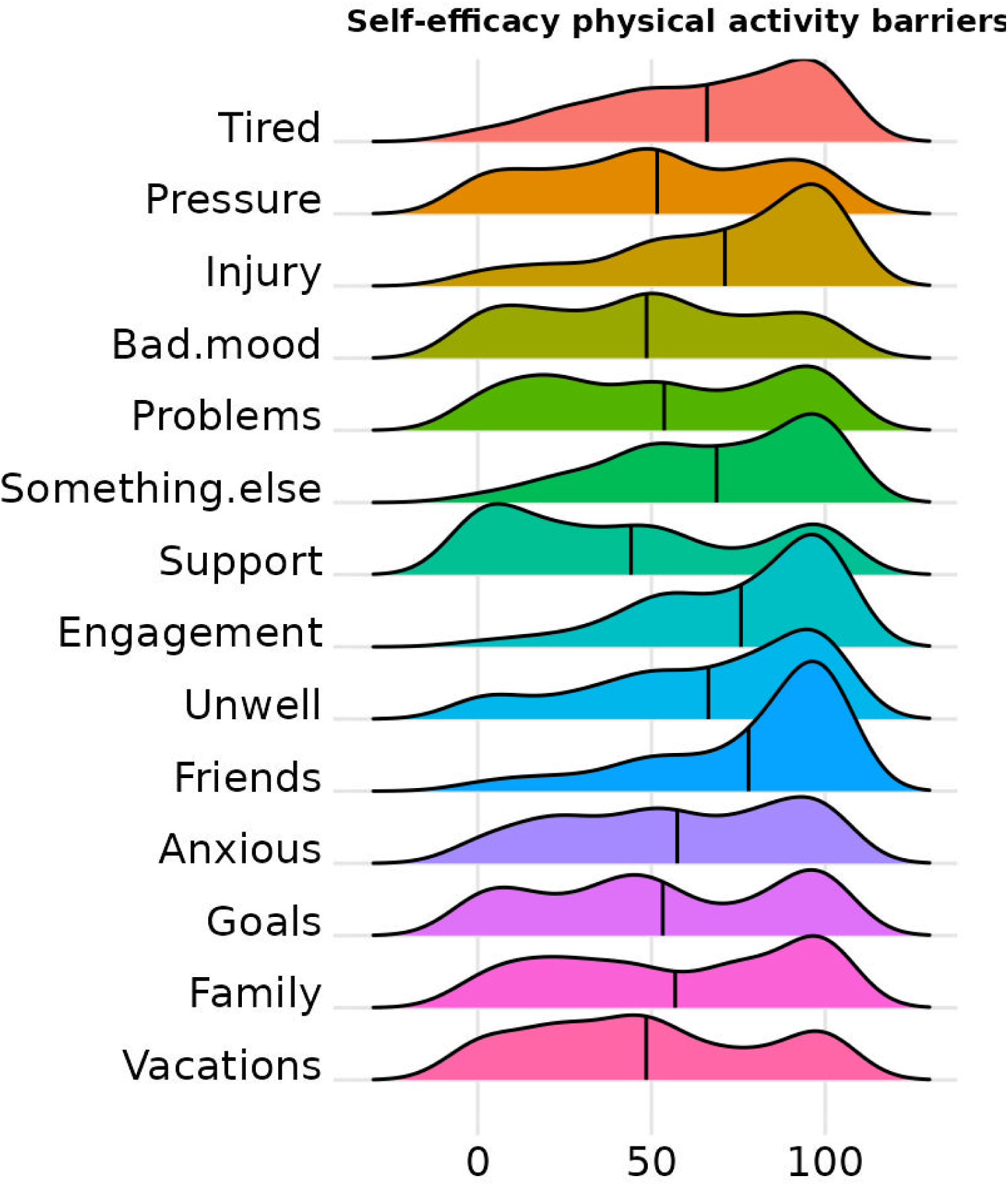
Self-efficacy to do PA when encountering barriers Note: The higher the mean indicator (vertical black line) on the abscissa, the greater the barrier. The different curves represent the distribution of data for each barrier. The barriers in order from the top are: “*Being tired*”, “*Feeling pressure at work*”, “*Recovering from an injury*”, “*Being in a bad mood*”, “*Having personal problems*”, “*Having more interesting things to do*”, “*Without the support of friends or family*”, “*Having other engagement*”, “*Feeling unwell*”, “*Having friends at home*”, “*Feeling anxious*”, “*Not reaching previously fixed training goals*”, “*Having family problems*”, “*During vacations*”.

### 3.3. Preferences

The detailed frequency to which each PA was declared as being among one’s preferred are represented in Figure 2. For detailed PA preferences according to body mass index, sex, country, education level, and age, see supplementary material (Figures S7-S11).

**Figure 2:**
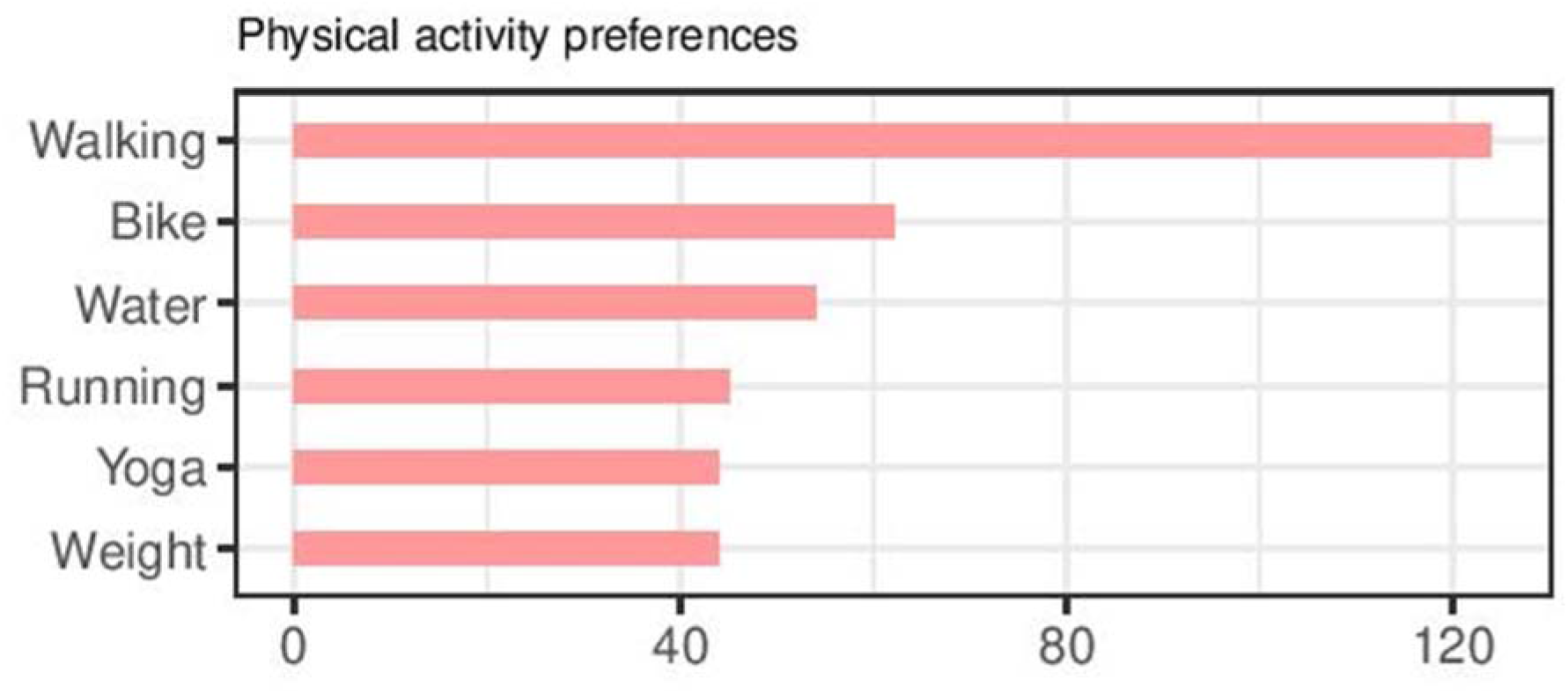
Most frequently preferred PA

The details for preferred modalities and contexts of PA are presented in Figure 3.

**Figure 3.**
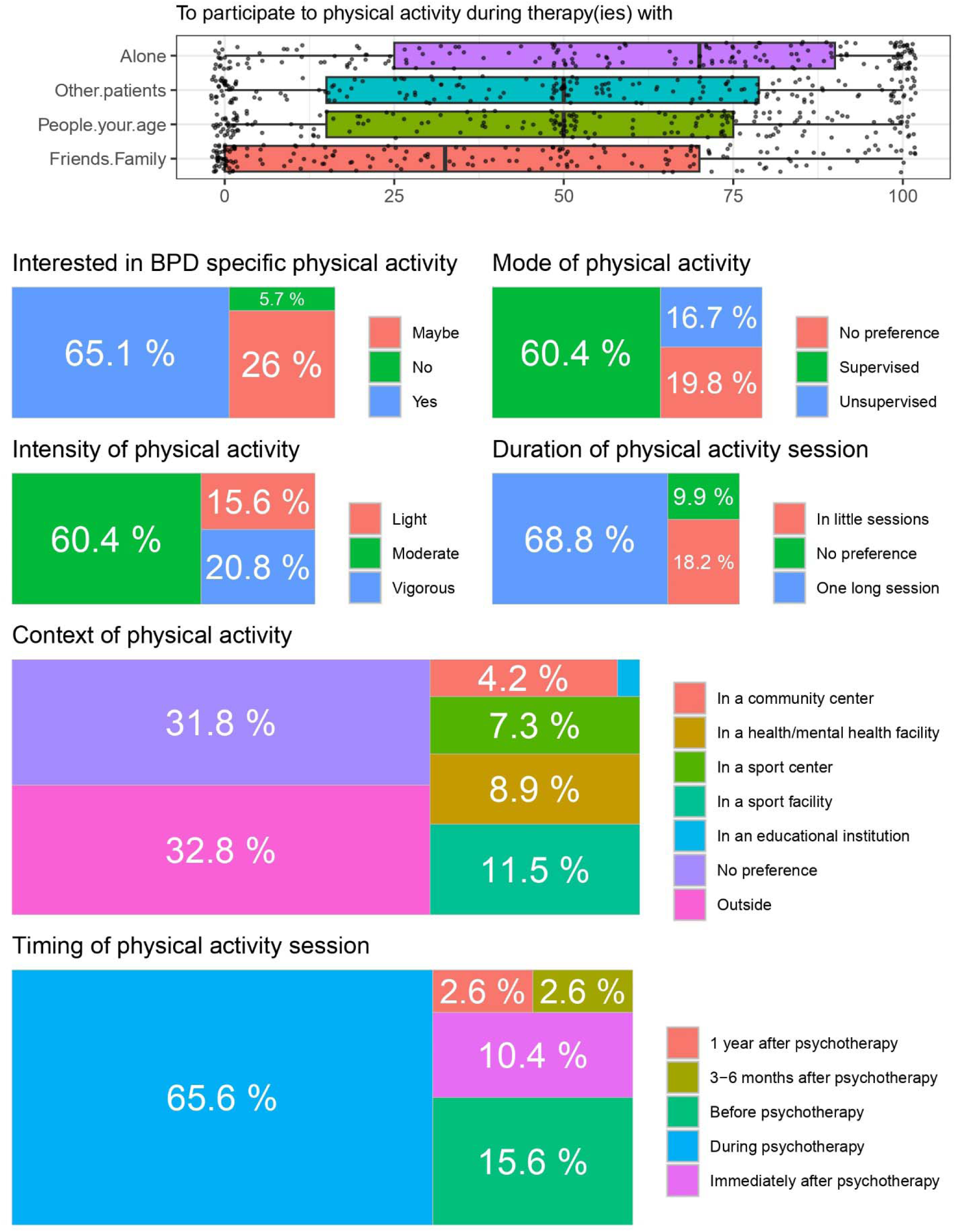
Preferred modalities and context of PA Note: BPD = borderline personality disorder; The sum of the % may not be equal to 100 due to missing data not included.

### 3.4. Advice

The details of the preferences regarding PA advice are presented in Figure 4.

**Figure 4.**
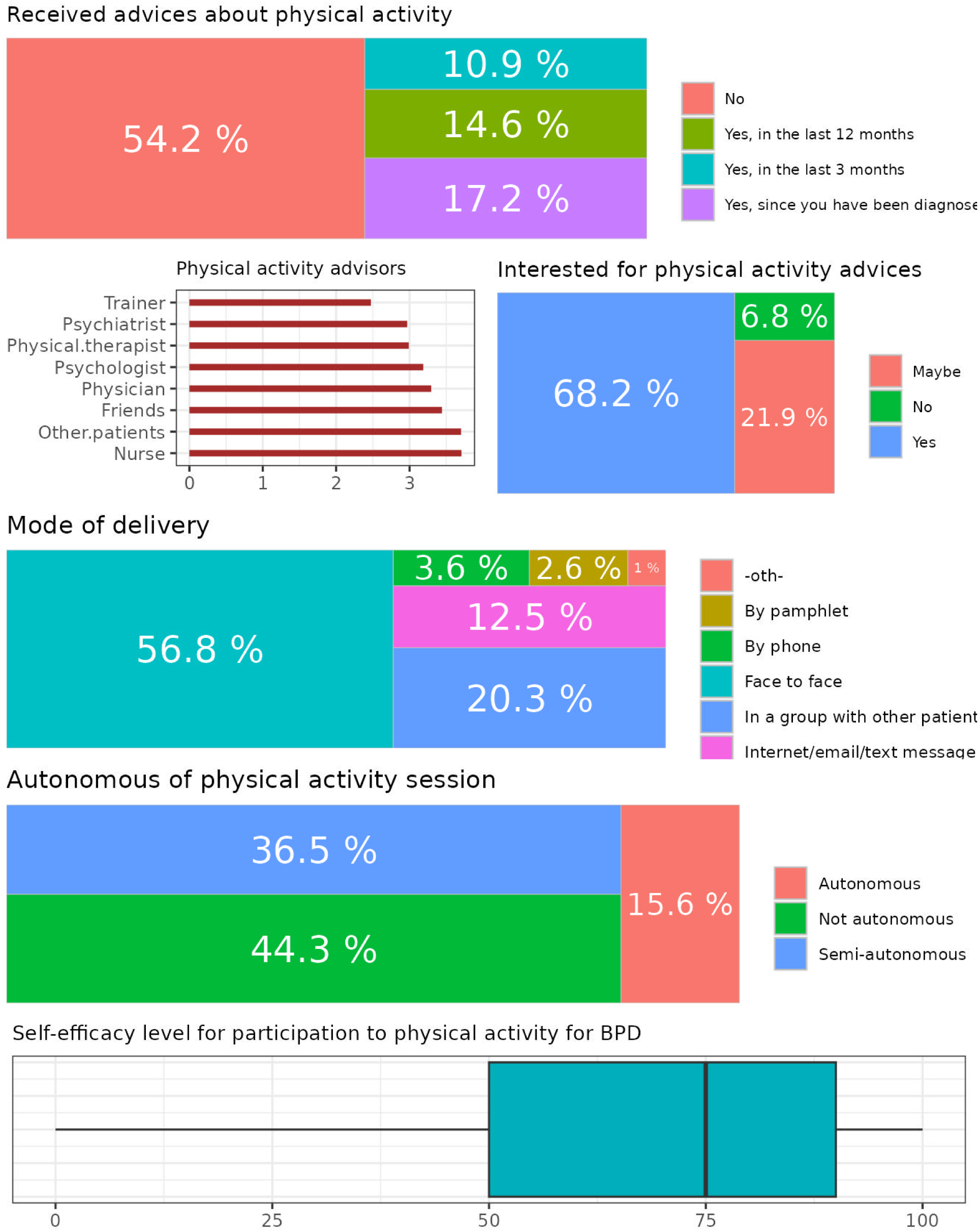
Preference regarding PA advice Note: For the PA advisor, the scores for each professional were calculated by asking participants to indicate which 3 professionals they prefer receiving advice from with the numbers 1 to 3 (1 = 3^rd^ choice; 2 = 2^nd^ choice; 3 = 1^st^ choice). All professional without number were attributed the number 0. The mean score for each professional (including 0s) were computed, reversed for clarity (making a mean score closer to 1 a higher preference for that professional) and reported here.

### 3.5. Correlates of PA

Seven univariate outliers have been excluded because they had an exceptional self-reported PA level (i.e., more than 3500 min per week). In this sample, the total level of PA is correlated to the age, the social level, the education level, the income, the body mass index, the level of BPD symptoms, having a tobacco use disorder, the level of depression, the number of psychotropic medications used and the self-efficacy to PA. For all the details see Table 2.

**Table 2:** Correlates to level of PA.

|  | Correlation with total self-reported PA |
| --- | --- |
| | $\beta$ (SD) |
| Age | -0.005*** (0.0003) |
| Perceived social status | 0.06*** (0.002) |
| University education | -0.23*** (0.007) |
| Income >40,000 | -0.31*** (0.007) |
| BMI | 0.006*** (0.0004) |
| BPD symptoms | 0.27*** (0.006) |
| Tobacco SUD | 0.38*** (0.007) |
| Alcohol SUD | 0.03*** (0.007) |
| Depression | -0.04*** (0.001) |
| Psychotropic medication | 0.09*** (0.003) |
| Self-efficacy | 0.01**; (0.002) |
| Observations | 180 |
| Adjusted R<sup>2</sup> | 0.12 |
| Log Likelihood | -66,656.41 |
| *Akaike information criterion* | 133,338.80 |

Note: BMI = body mass index; BPD = borderline personality disorder; SUD = substance use disorder.
\*\*\* $p < 0,01$

## 4. Discussion

This is the first study to describe the PA level, preferences, barriers and correlates in a sample of adults with BPD. Although online studies pose many challenges, they are efficient and doable in adults with BPD (DeShong & Tucker, 2019; Lawn & McMahon, 2015).

### 4.1. Level of PA

First, we observe a relatively high level of PA in this sample. With a mean of more than 600 minutes weekly, participants are on average more active than the general population (about 340 minutes weekly; Colley et al., 2018) and individuals with mental disorders (about 270 minutes weekly; Vancampfort et al., 2017). With a little over 60%, this sample presents a similar proportion of active individuals as the general population (about 60%; Colley et al., 2018) but a bigger proportion than individuals with other mental disorders (about 45%; Vancampfort et al., 2017). These data are surprising and might indicate that the mean PA level is skewed by extreme individuals with large PA volume. However, the present data are difficult to put in perspective because of the absence of studies reporting PA level in this population (St-Amour et al., 2023).

### 4.2. Barriers

The main barriers to PA reported in this study, as measured by the lowest reported self-efficacy to overcome them, are having friends over, having other engagements, recovering from an injury, having something else more interesting to do and feeling too tired. In comparison, individuals with severe mental disorders or substance use disorder reported lack of motivation, fatigue, having no one to engage in PA with, not having enough energy, would not be able to keep up, and lack of financial resources as barriers to PA (Abrantes et al., 2011; Romain, Longpré-Poirier, et al., 2020). The barriers reported here were then somewhat different from those reported in other populations with mental disorders with the exception of lack of energy/feeling tired. Surprisingly, lacking support or not having anyone to engage in PA with was the least important barrier in adults with BPD but was among the main barriers in populations with other mental disorders (Abrantes et al., 2011; Romain, Longpré-Poirier, et al., 2020). This difference might be explained by the relational nature of the personality disorder. Indeed, one of the main characteristics of BPD is the difficulty in maintaining interpersonal relationships (Euler et al., 2019). These difficulties might lead individuals with BPD to prefer engaging in PA alone and therefore not considering lacking support or not having someone to do PA with as a major barrier to PA. Moreover, individuals with BPD tend to be more impulsive and to prefer quick smaller rewards than delayed larger rewards than healthy individuals and those with other mental disorders (Gunderson et al., 2018). Therefore, they might be tempted to engage in quickly rewarding activities rather than PA. However, more studies are needed to thoroughly understand this difference.

### 4.3. Preferences

The most frequently reported favourite PAs are somewhat consistent with the findings from populations with other mental disorders (Abrantes et al., 2011; Romain, Longpré-Poirier, et al., 2020). Indeed, walking is the preferred PA regardless of the studied population (Abrantes et al., 2011; Romain, Longpré-Poirier, et al., 2020). The clear preference of walking as a PA might be attributed to it being practical, self-paced and controlled, inexpensive, and not needing a lot of resources (Abrantes et al., 2011).

As mentioned before, participants in this study clearly prefer engaging in PA alone than with other people. This result differs greatly from what is seen in other populations with mental disorder having mostly no preference for doing PA alone or in groups. This difference might also be explained by the interpersonal difficulties experienced by individuals with BPD (Euler et al., 2019).

In this sample, most participants prefer supervised PA compared to unsupervised PA. This result is also surprising and in opposition to the findings observed in other populations. Indeed, in populations with other mental disorders, there is either a clear preference for unsupervised PA or no clear preference regarding supervision (Abrantes et al., 2011; Romain, Longpré-Poirier, et al., 2020). This result is unexpected considering previously observed results and interpersonal difficulties of individuals with BPD. However, this difference might be explained by the perception of the different relationships in play. Individuals with BPD tend to feel closer from those peripheral to their social network and farther from the more central individuals (Beeney et al., 2018). Therefore, they might feel closer to an outside individual like a kinesiologist or physical therapist supervising their PA session but would not want to share these moments with friends or relatives.

### 4.4. Advice

An overwhelming majority of participants declared being interested or maybe interested in receiving PA advice. However, a majority also declared not having received PA advice. Since all participants reportedly received BPD diagnosis from a healthcare professional, this lack of advice regarding PA is alarming considering two evidence-based BPD treatments suggest PA in their official guidelines (Blum et al., 2008; Linehan, 2014). Moreover, participants declared trusting healthcare professionals in giving them advice about PA, but preferred trainers, psychiatrists and physical therapists. It would therefore be important to include kinesiologists and PA professionals in multidisciplinary teams taking care of individuals with BPD. Future research may look at different PA promotion interventions in this population specifically (motivational interviews, messaging, group intervention, etc.).

### 4.5. Correlates of PA

In this study, sociodemographic and clinical variables have some opposite correlation to PA level with what is observed in previous systematic reviews (Vancampfort et al., 2012). Indeed, age, education level, and household income are negatively correlated and social status is positively correlated to PA level. Also, body mass index, BPD symptoms, tobacco use disorder, alcohol use disorder and number of psychotropic medications are positively correlated. Higher levels of depression is associated with lower PA level. Finally, self-efficacy level is also positively linked to PA level which is also observed in a recent meta-analysis (Cabassa et al., 2020). Among these correlations, the most surprising are those with the education level, the household income, the level of BPD symptoms, the tobacco and alcohol use disorder, and the number of psychotropic medications. The association between lower income and education level, and higher PA level in the present sample might be explained by a greater proportion of work-related PA in individuals with lower income and education level (Prince et al., 2020). Moreover, work-related PA is more strongly correlated with the total PA volume than leisure time PA and most studies reporting PA level only report leisure time PA (Vancampfort et al., 2012). Health-related surprising correlates (body mass index, BPD symptoms, substance use disorders and medication) might be explained by the nature of psychiatric comorbid disorders in our participants (St-Amour, Hains-Monfette, et al., 2021; Vancampfort et al., 2012).

### 4.6. Limitations

However, this study suffers from some limitations. First, BPD diagnosis was self-reported making it hard to ensure this sample is composed solely of adults with BPD. This sample is also small considering the observational design adopted. Second, this study could have attracted more active than inactive participants due to its theme (i.e., PA) subjecting it to recruitment bias and increasing PA levels artificially. Future studies could use representative national surveys reporting both BPD diagnosis and PA. However, to our knowledge, no such survey is available at the moment. Moreover, the unequal recruitment between countries biases the results with an over-representation of Canada and France limiting the generalizability in other countries. The online nature of this survey also poses some limitations. Indeed, online surveys are only accessible to those with access to a computer and sufficient informatics literacy to complete them. In addition, the cross-sectional design of this study prevents us from establishing causality relations between PA and its correlates. Finally, participants may have developed questionnaire fatigue because of the length of the survey, but this was not measured nor analyzed.

## 5. Conclusion

To our knowledge, this is the first study analyzing PA level, preferences, barriers, and correlates in adults with BPD. This information is primordial in developing future studies analyzing PA’s medium- to long-term effect in adults with BPD. With treatment guidelines already suggesting the use of PA (Blum et al., 2008; Linehan, 2014), and indirect evidence indicating potential benefits of PA in alleviating BPD symptoms (Mehren et al., 2020), there is an urgent need for data analyzing the effect of PA on the different components of BPD to test this hypothesis. Pending future results, PA could be used by healthcare professionals (mostly kinesiologists, psychiatrists, and psychologists) to treat or alleviate comorbid disorders in this population. In doing so, they should base their intervention on the present barriers and preferences (i.e., walking, individual intervention, supervised sessions, outdoor, etc.) to ensure greater adherence (Romain, Bernard, et al., 2020). Future studies should also use nationally representative surveys to answer these questions and aim to compare these results with those from individuals with other metal disorders.

## Data Availability

All data produced are available online at

https://osf.io/u5sgj/

