## Supplementary material for "Pattern, preferences, barriers, and correlates of self-reported physical activity in adults with borderline personality disorder: An online survey in western countries": Figure S

Table of Content

### Figure S1: physical activity level according to body mass index, sex, level of education, age, and country

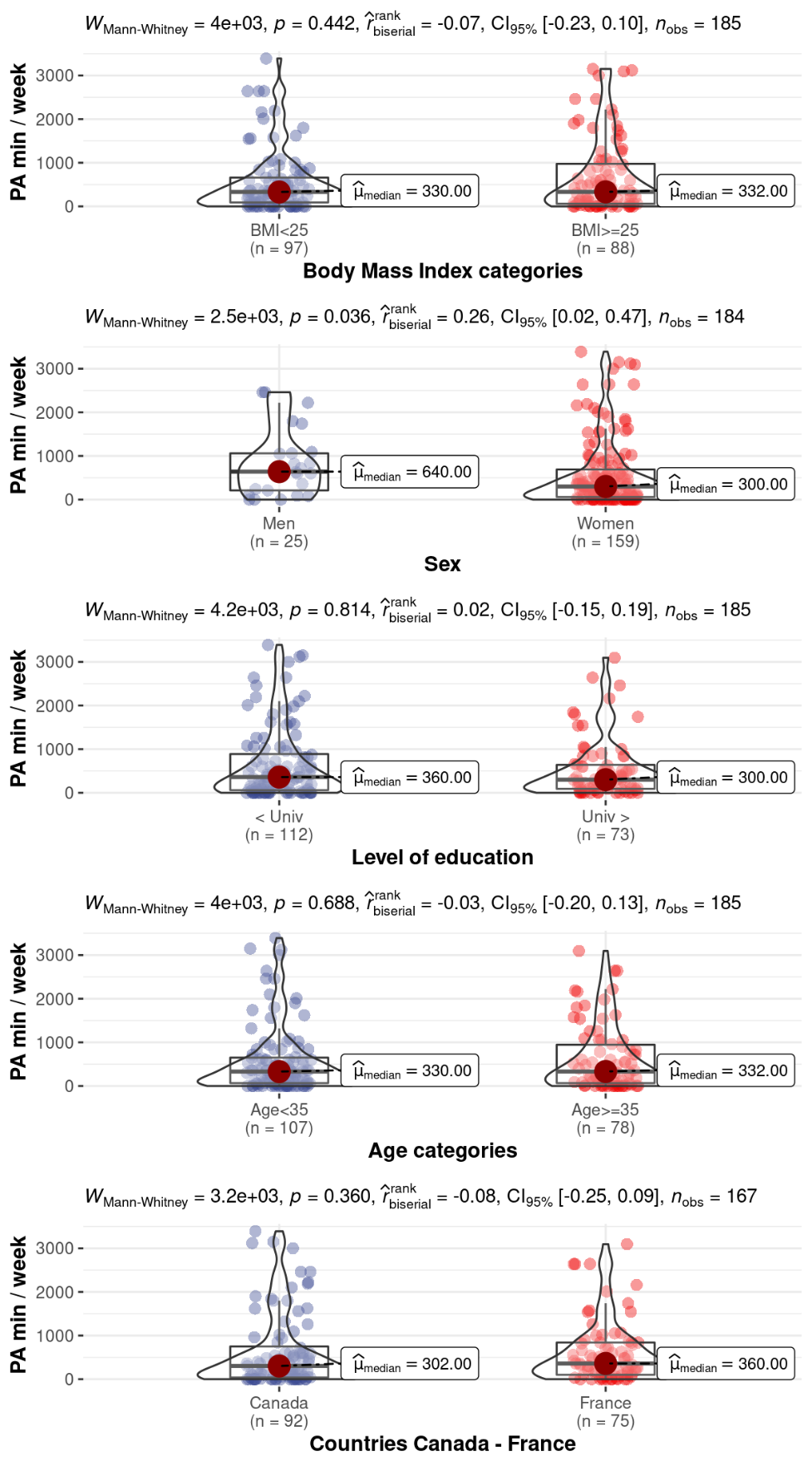

Note : PA = Physical activity.

### Figure S2: Self-efficacy to do physical activity when encountering barrier according to age

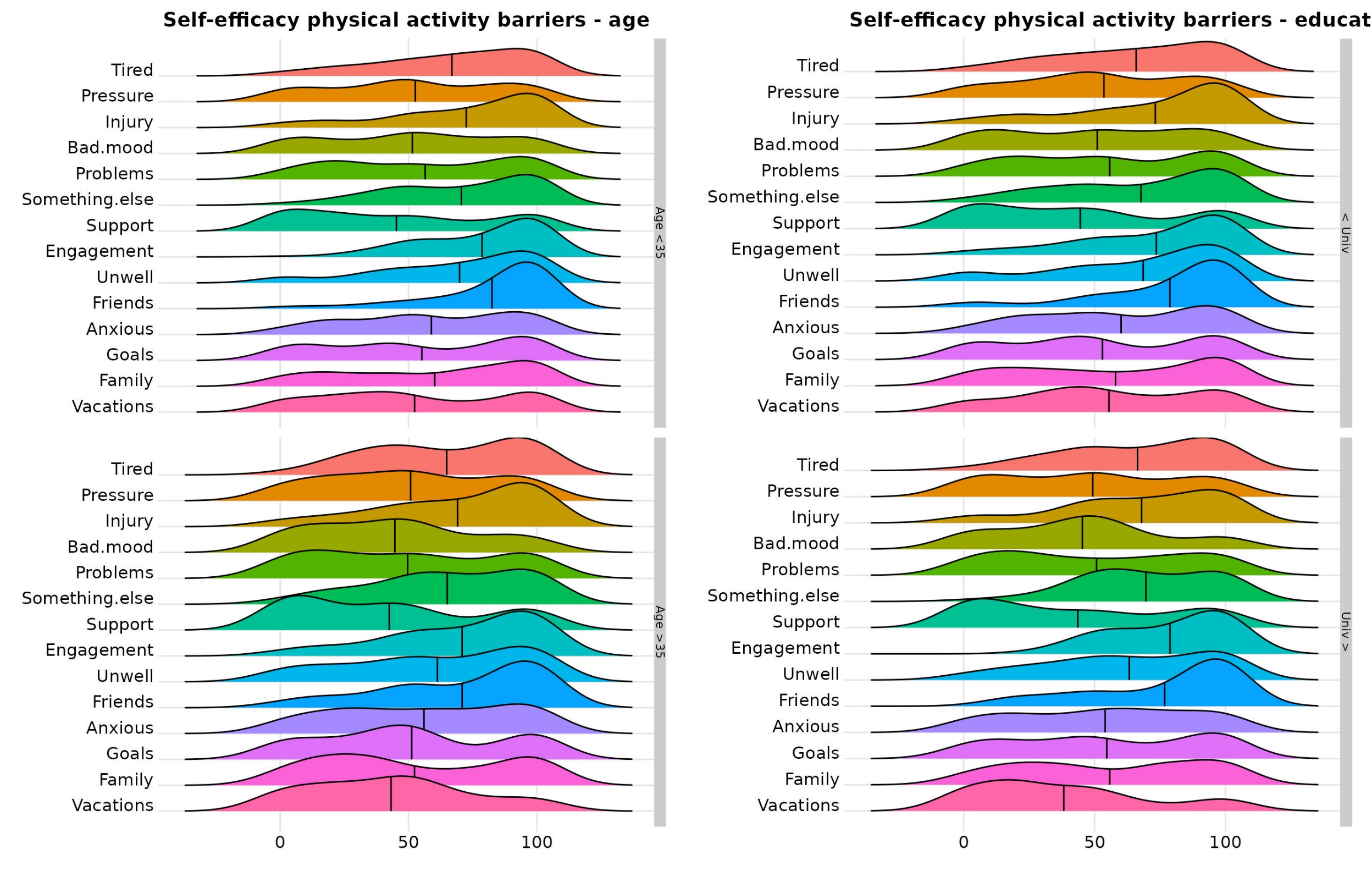

Note: The higher the mean indicator (vertical black line), the greater the barrier. The barriers in order from the top are: “*Being tired*”, “*Feeling pressure at work*”, “*Recovering from an injury*”, “*Being in a bad mood*”, “*Having personal problems*”, “*Having more interesting things to do*”, “*Without the support of friends or family*”, “*Having other engagement*”, “*Feeling unwell*”, “*Having friends at home*”, “*Feeling anxious*”, “*Not reaching previously fixed training goals*”, “*Having family problems*”, “*During vacations*”.

### Figure S3: Self-efficacy to do physical activity when encountering barrier according to education level

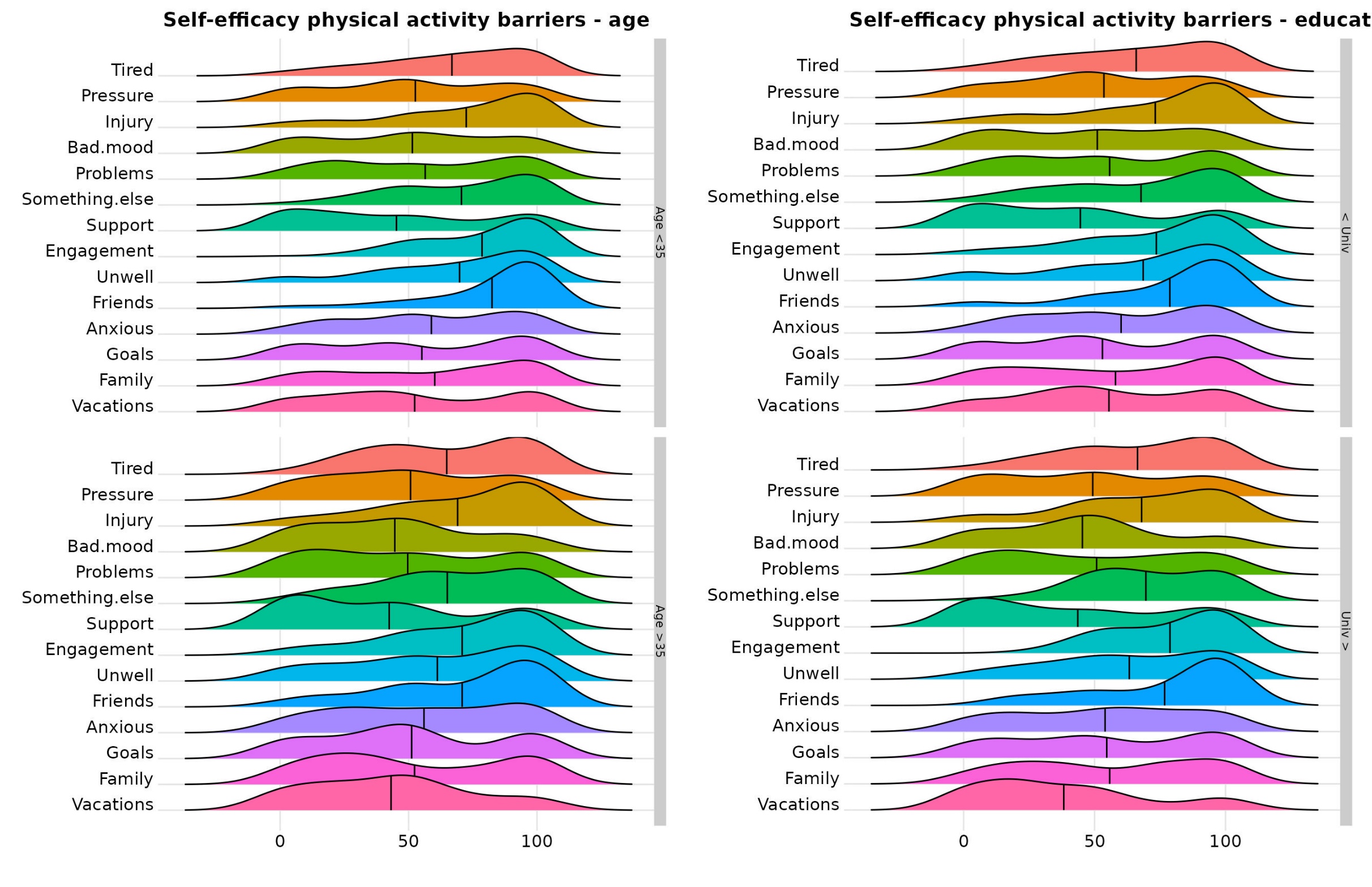

Note: The higher the mean indicator (vertical black line), the greater the barrier. The barriers in order from the top are: “*Being tired*”, “*Feeling pressure at work*”, “*Recovering from an injury*”, “*Being in a bad mood*”, “*Having personal problems*”, “*Having more interesting things to do*”, “*Without the support of friends or family*”, “*Having other engagement*”, “*Feeling unwell*”, “*Having friends at home*”, “*Feeling anxious*”, “*Not reaching previously fixed training goals*”, “*Having family problems*”, “*During vacations*”.

### Figure S4: Self-efficacy to do physical activity when encountering barrier according to sex

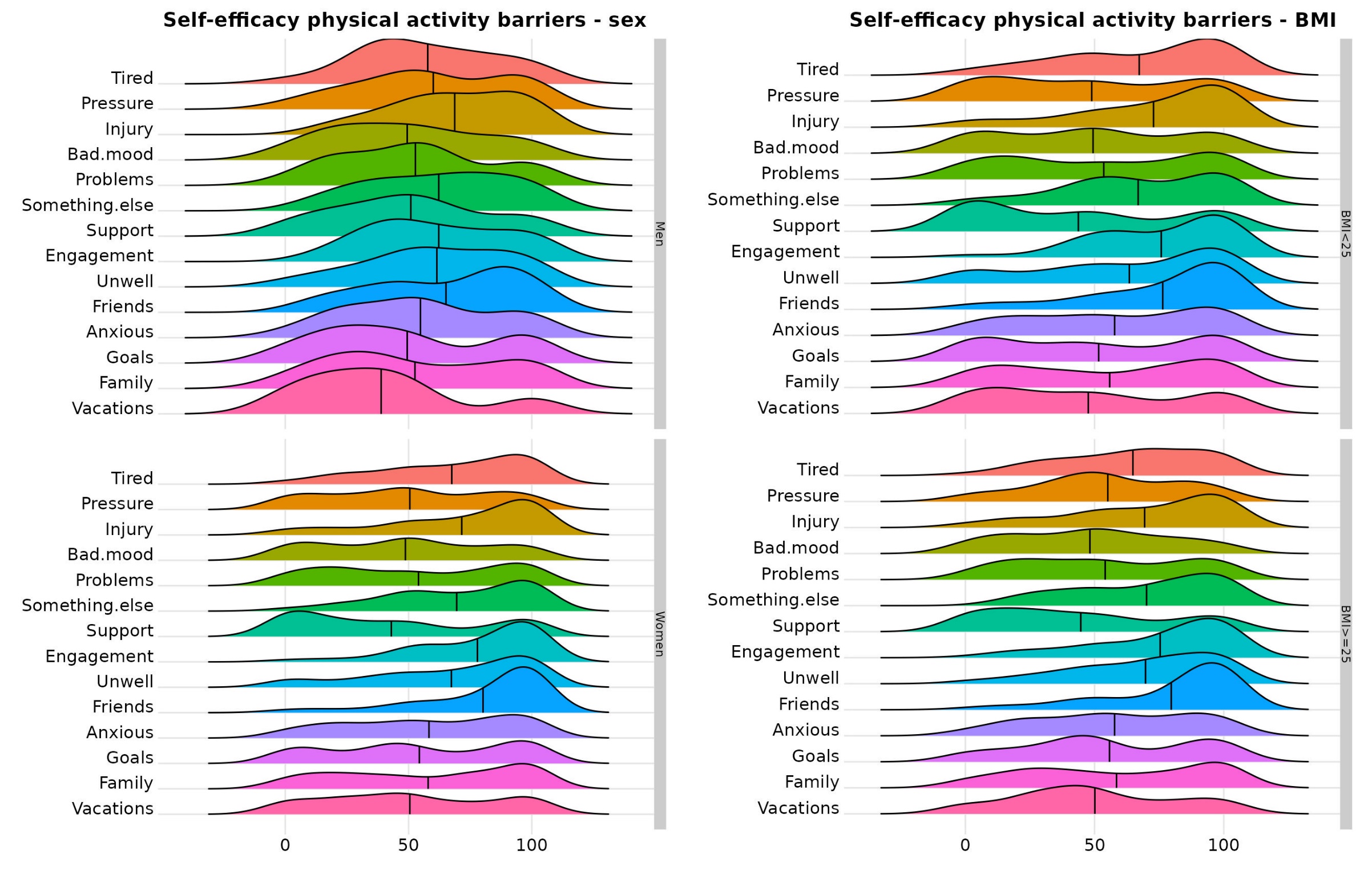

Note: The higher the mean indicator (vertical black line), the greater the barrier. The barriers in order from the top are: “*Being tired*”, “*Feeling pressure at work*”, “*Recovering from an injury*”, “*Being in a bad mood*”, “*Having personal problems*”, “*Having more interesting things to do*”, “*Without the support of friends or family*”, “*Having other engagement*”, “*Feeling unwell*”, “*Having friends at home*”, “*Feeling anxious*”, “*Not reaching previously fixed training goals*”, “*Having family problems*”, “*During vacations*”.

### Figure S5: Self-efficacy to do physical activity when encountering barrier according to body mass index

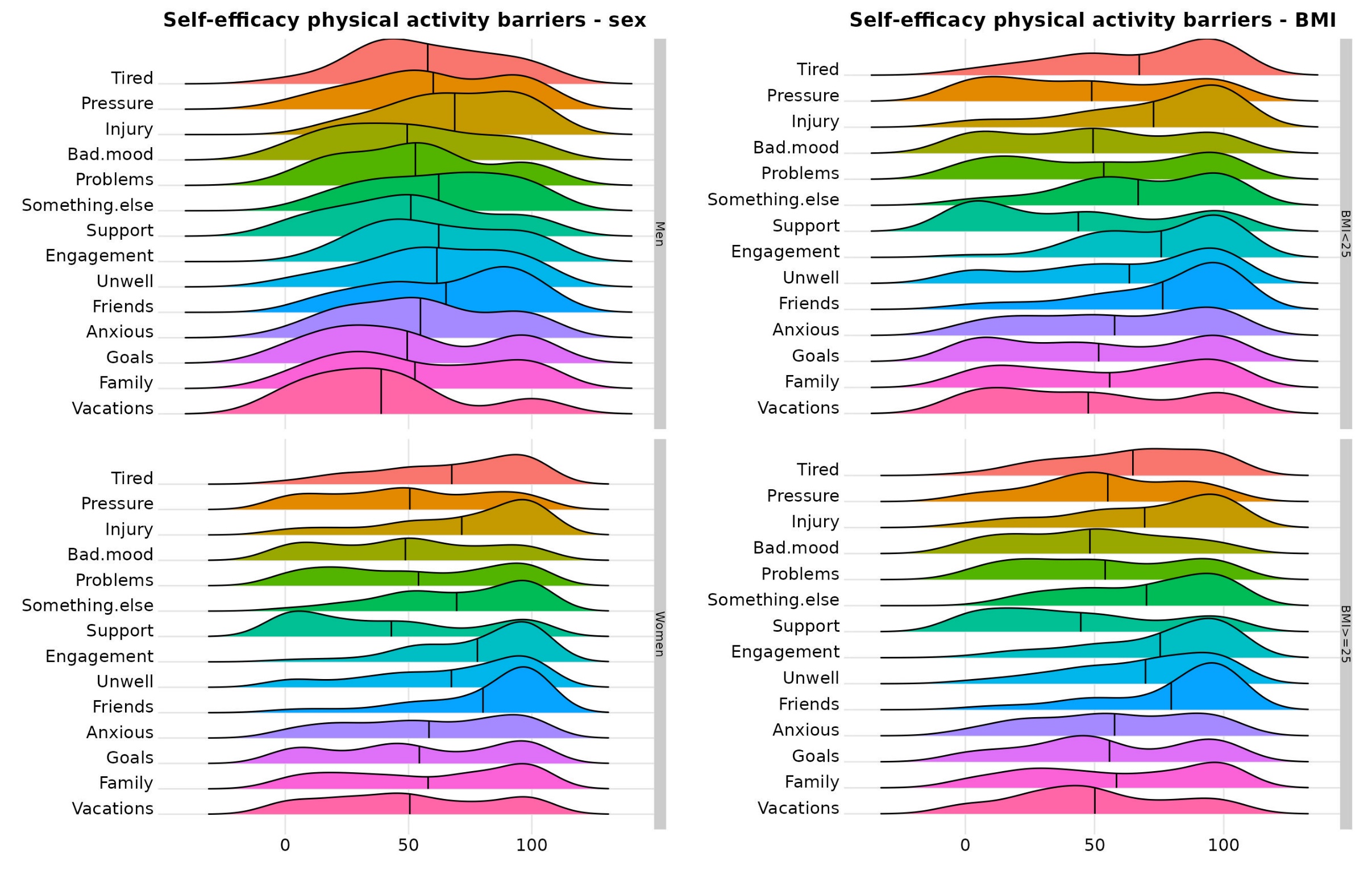

Note: BMI = Body mass index. The higher the mean indicator (vertical black line), the greater the barrier. The barriers in order from the top are: “*Being tired*”, “*Feeling pressure at work*”, “*Recovering from an injury*”, “*Being in a bad mood*”, “*Having personal problems*”, “*Having more interesting things to do*”, “*Without the support of friends or family*”, “*Having other engagement*”, “*Feeling unwell*”, “*Having friends at home*”, “*Feeling anxious*”, “*Not reaching previously fixed training goals*”, “*Having family problems*”, “*During vacations*”.

### Figure S6: Self-efficacy to do physical activity when encountering barrier according to country

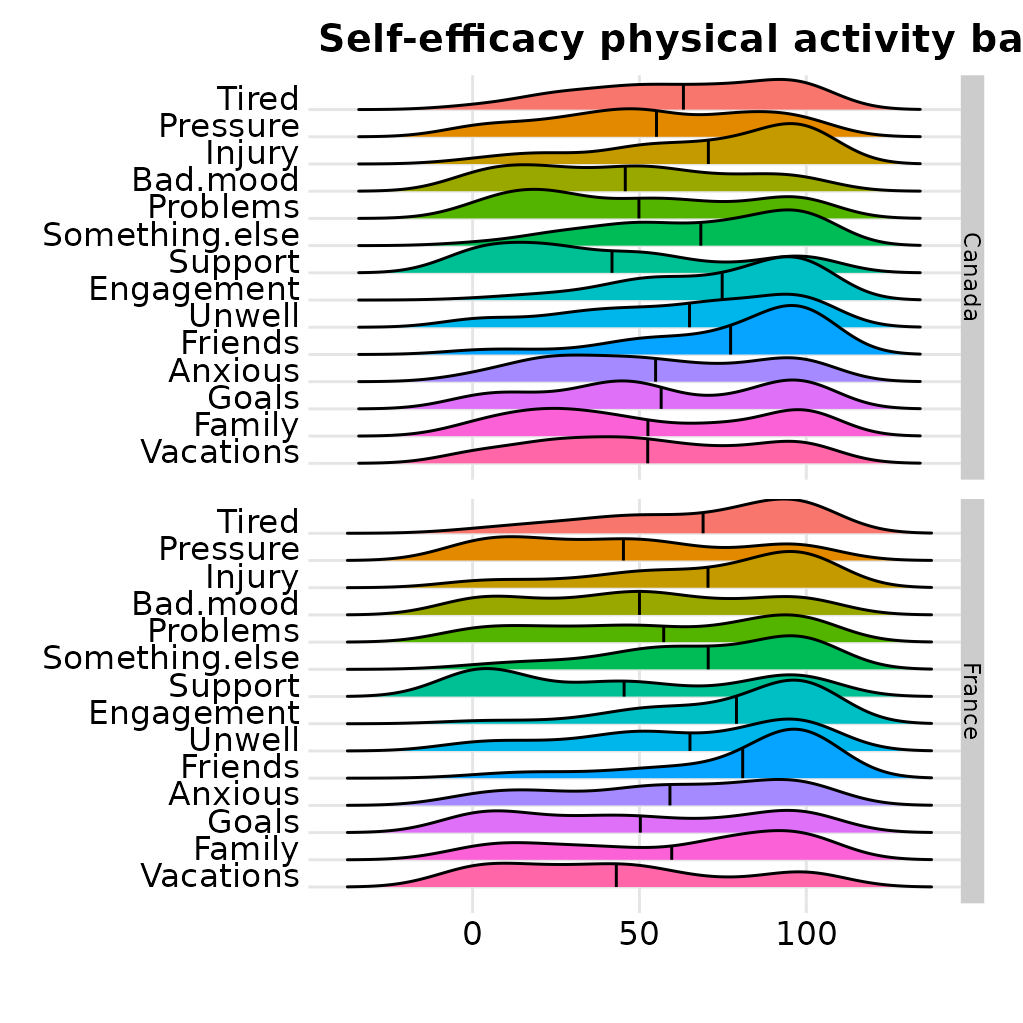

Note: The higher the mean indicator (vertical black line), the greater the barrier. The barriers in order from the top are: “*Being tired*”, “*Feeling pressure at work*”, “*Recovering from an injury*”, “*Being in a bad mood*”, “*Having personal problems*”, “*Having more interesting things to do*”, “*Without the support of friends or family*”, “*Having other engagement*”, “*Feeling unwell*”, “*Having friends at home*”, “*Feeling anxious*”, “*Not reaching previously fixed training goals*”, “*Having family problems*”, “*During vacations*”.

### Figure S7: Physical activity preferences according to body mass index

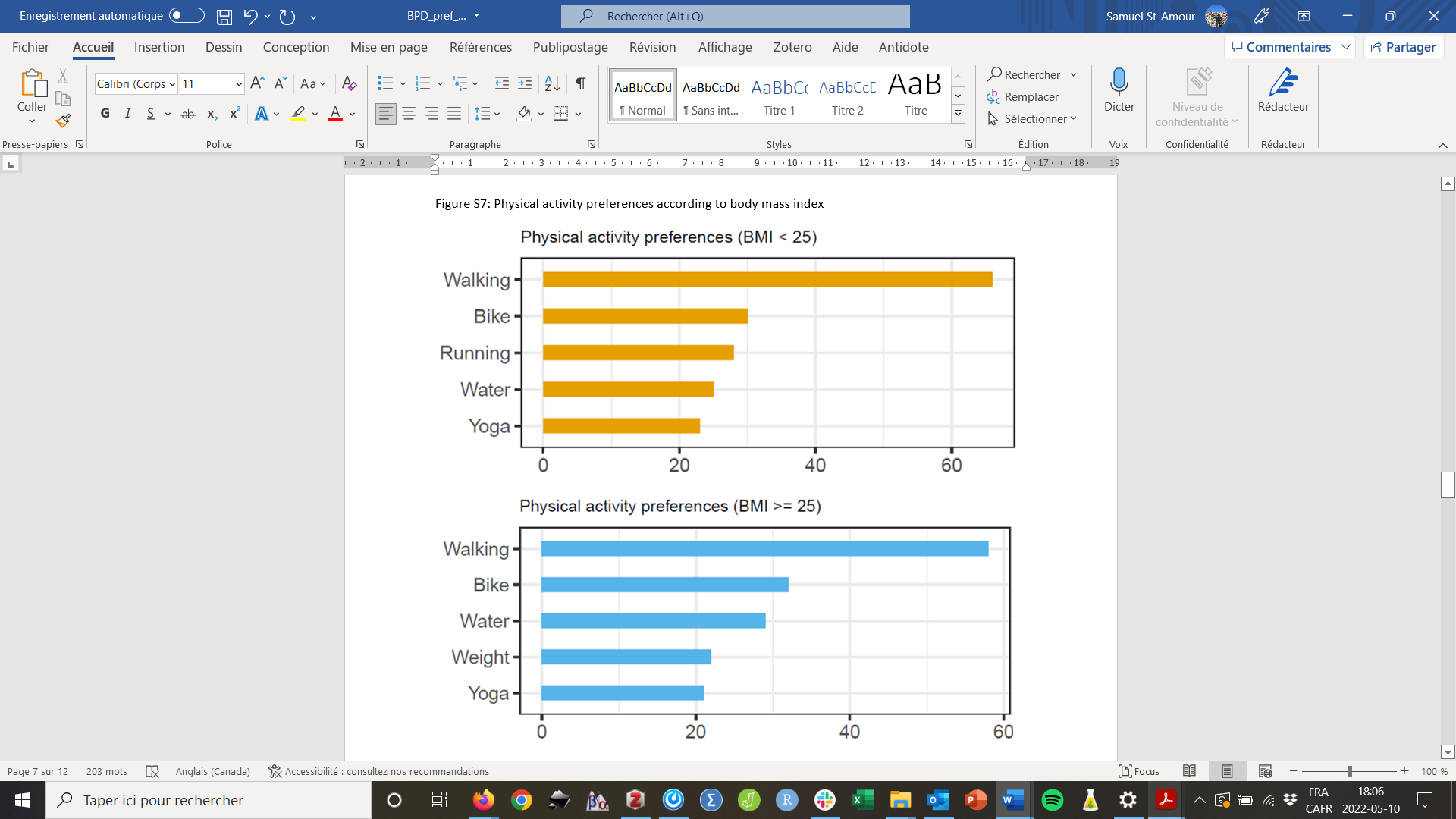

### Figure S8: Physical activity preferences according to sex

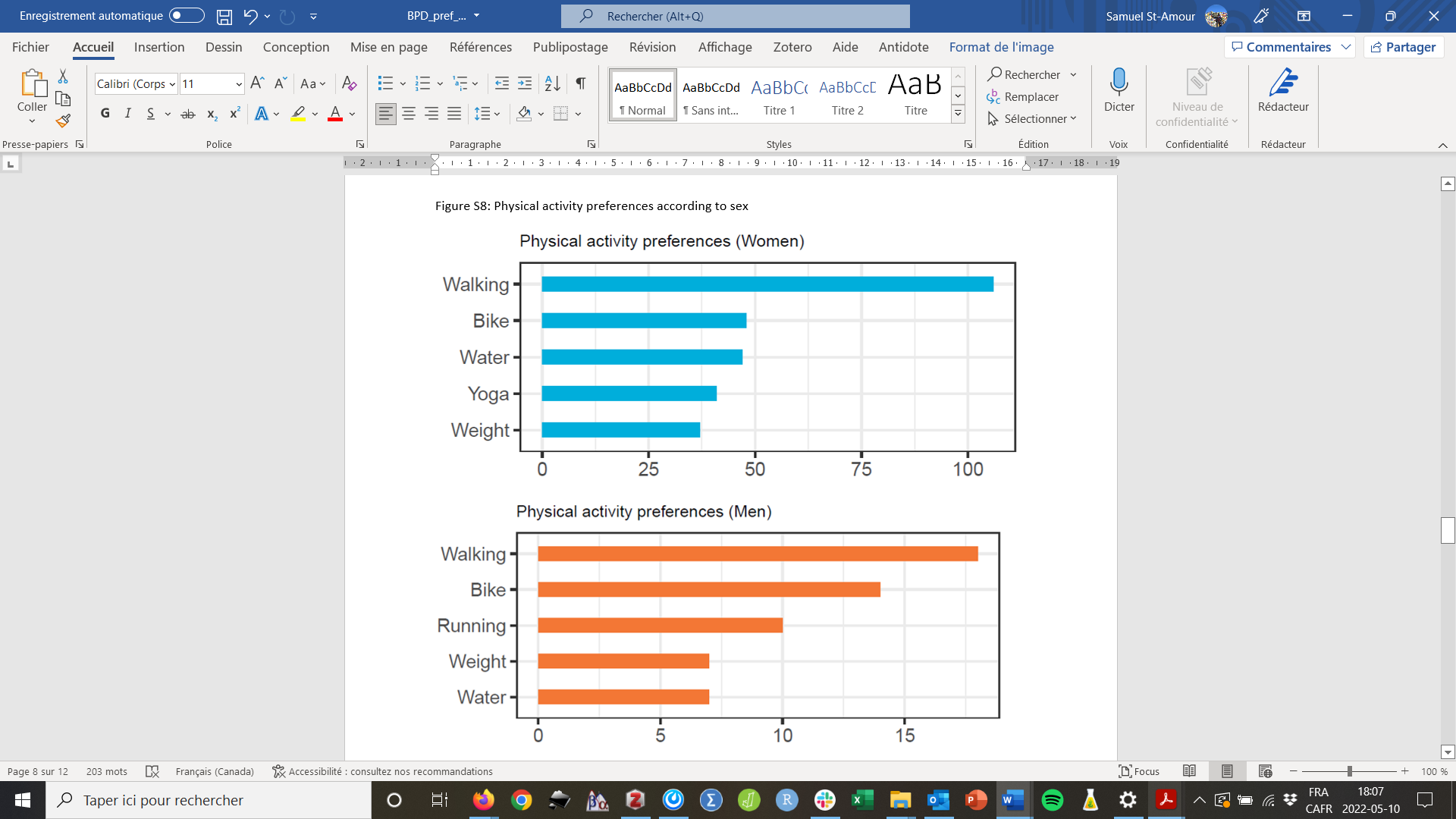

### Figure S9: Physical activity preferences according to country

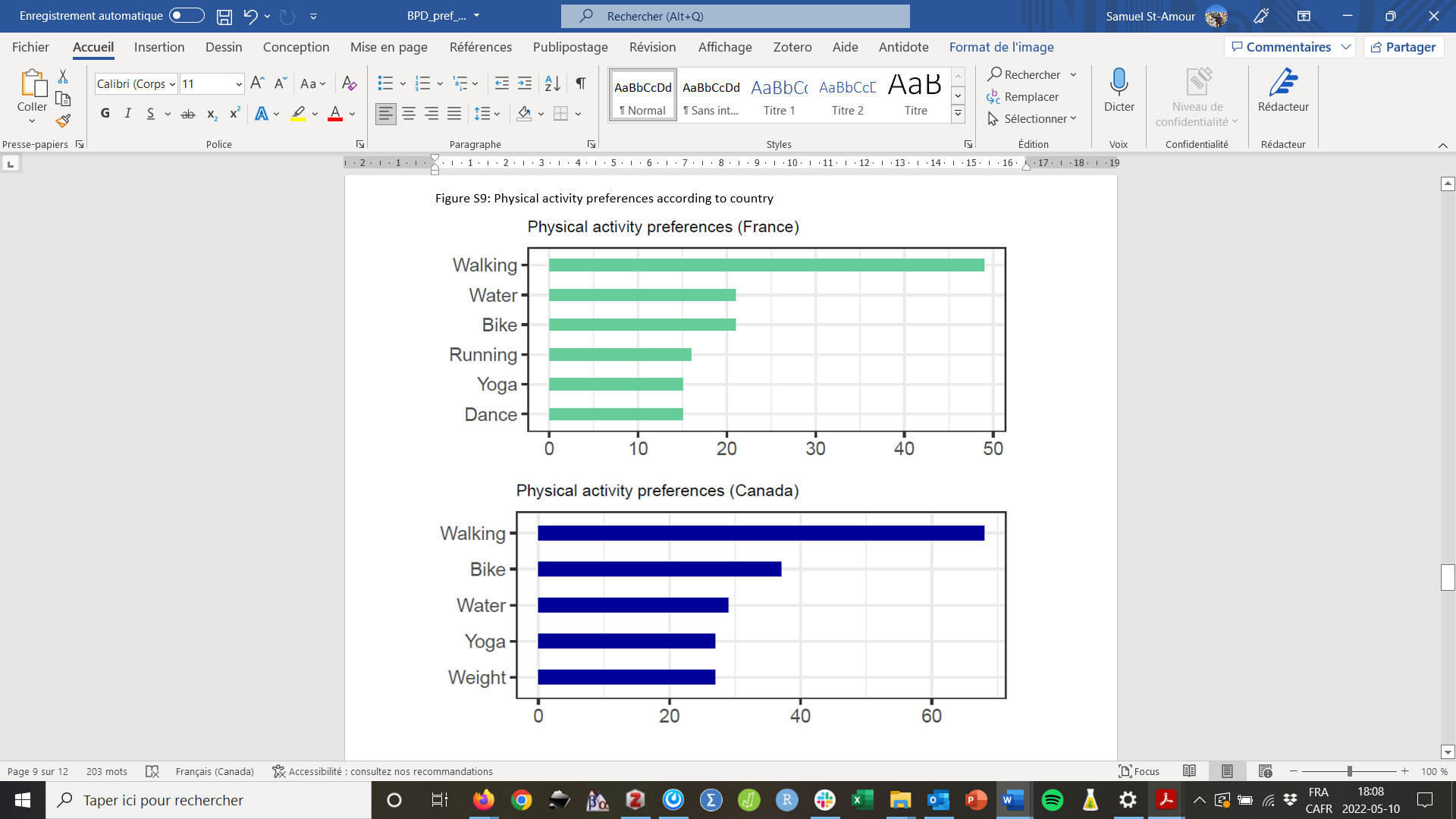

### Figure S10: Physical activity preferences according to education level

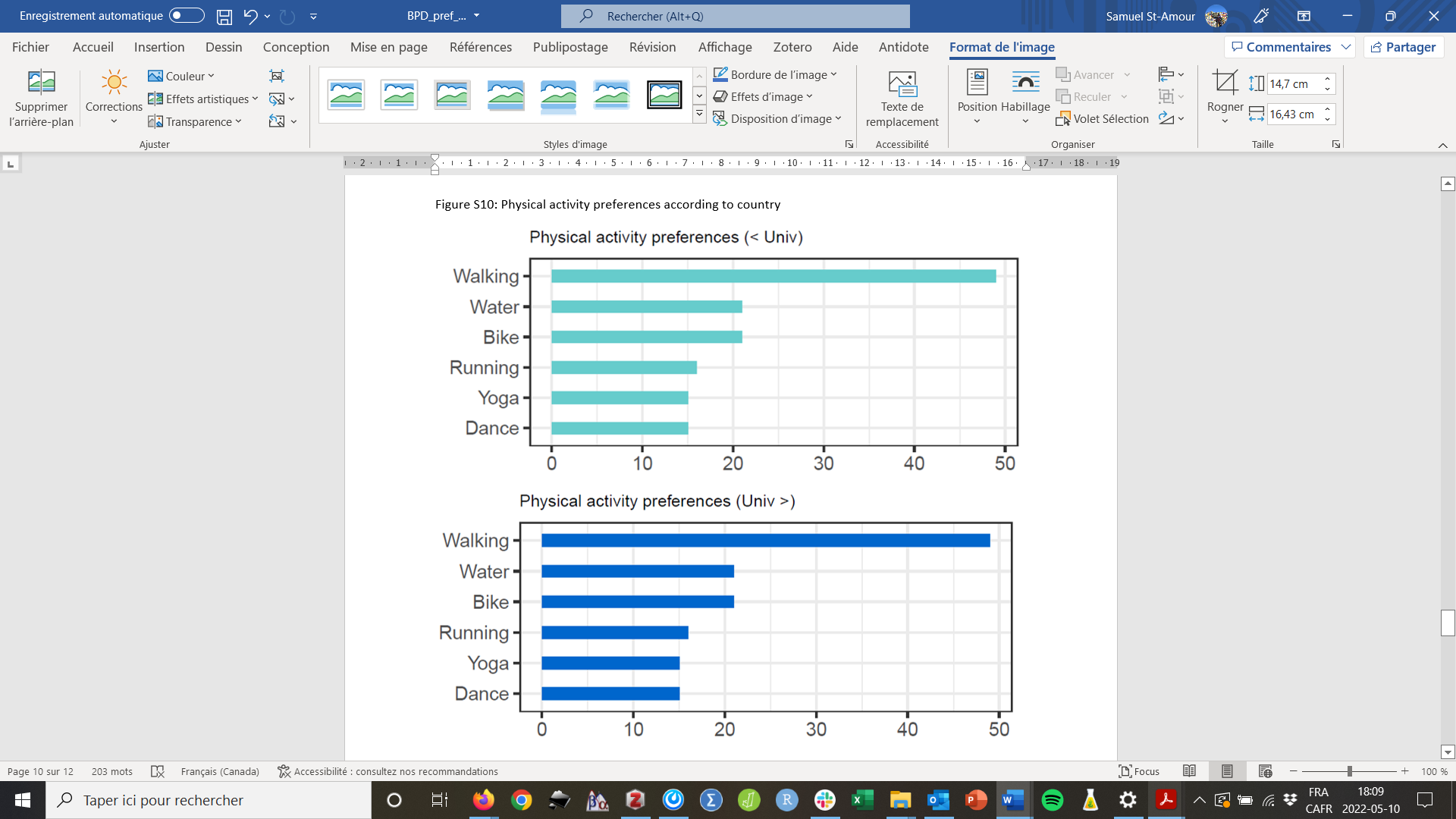

### Figure S11: Physical activity preferences according to age

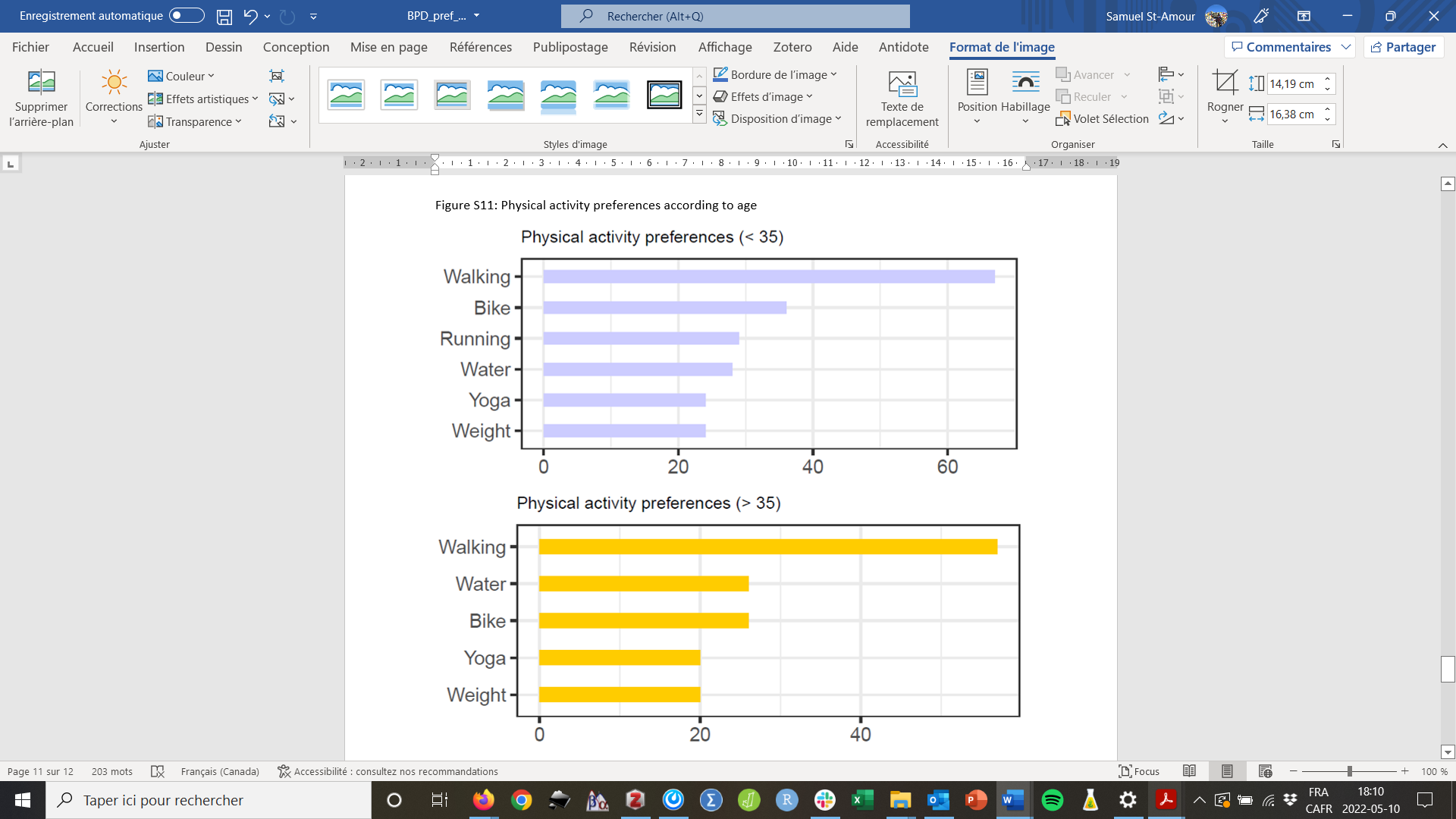
